# Vision-Based Automated Severity Rating of REM Sleep Behavior Disorder: From Heuristic Features to Foundation Models

**DOI:** 10.64898/2026.07.02.26356954

**Authors:** Jeongyeon Hwang, Nicolas van Pesch, Brinda Raval, Mohamed Abdelfattah, Amene Gafsi, Amelia Bertolaso, Kang Hyun Ryu, Salonee Marwaha, Oliver Sum-Ping, Matteo Cesari, Ambra Stefani, Andreas Brink-Kjaer, Emmanuel Mignot, Alexandre Alahi, Emmanuel During

## Abstract

Automated severity rating of rapid eye movement (REM) sleep behavior disorder (RBD) movements would enable multi-night monitoring to detect potentially injurious behaviors and provide objective endpoints for clinical trials. We compared a heuristic classifier using optical flow-derived features against V-JEPA2, a self-supervised video foundation model, for clip-level severity classification (3329 mild versus 284 moderate-to-severe) of in-laboratory video-polysomnography infrared recordings in 86 isolated RBD patients. V-JEPA2 with checkpoint fine-tuning and maximum optical flow-based frame sampling achieved the best performance across both evaluation conditions — Macro F1 of 0.76 and 93% accuracy in the clip-level split, and 0.68 and 85% in the patient-level split — outperforming heuristic and domain-specific pretrained models. Clip duration was the dominant heuristic predictor. Whole-night severity scores preserved patient-level ordering despite systematic overestimation, with V-JEPA2 achieving a mean absolute error of 25% versus 52% for the heuristic classifier. These findings establish a foundation for objective, home-deployable monitoring of RBD severity.

## BACKGROUND

Rapid eye movement (REM) sleep behavior disorder (RBD) is a parasomnia characterized by abnormal motor disinhibition resulting from the loss of normal muscle atonia during REM sleep^1^ and is associated with a high risk of injury.^1^ RBD affects approximately 2% of the adult population aged 60 years and older, representing nearly 25 million people worldwide. In most patients, RBD is associated with an underlying alpha-synucleinopathy, including Parkinson’s disease and dementia with Lewy bodies, and manifestations often emerge during the prodromal phase of these neurodegenerative diseases.^2–4^

Clinically, RBD is characterized by vivid, action-filled, often unpleasant dreams that can lead to potentially injurious behaviors for patients and bed partners including punching, kicking, or falling out of bed.^2–5^ Symptomatic treatment options remain limited, largely confined to melatonin and clonazepam. Melatonin demonstrates variable efficacy, while clonazepam is associated with tolerability concerns, especially in older or medically complex patients.^6^ Better therapies are needed; however, clinical trials have been relatively scarce, and most controlled studies have failed to demonstrate superiority of active drugs over placebo. A key contributor to this limitation is the absence of well-defined, reliable, and objective endpoints for evaluating treatment efficacy.^7^

To date, most trials have relied primarily on patient- and partner-reported outcomes, which are susceptible to recall bias, expectation effects, and the high night-to-night variability of RBD symptoms.^8^ Recently, the International RBD Study Group (IRBDSG) set a research agenda for the development of safer and more effective therapies and recommended the use of video for objective assessment of symptom severity using a 3-point scale for mild, moderate, and severe motor behaviors.^9^ Severe movements are defined as forceful, vigorous, or violent behaviors that could severely injure the patient or bed partner (e.g., kicking or punching), while mild movements are defined as those with no risk of injury, and moderate movements represent an intermediate category. A composite whole-night severity score can then be calculated by weighting mild, moderate, and severe movements as 1, 5, and 10 points, respectively.^9,10^ This framework, however, depends on expert manual review, which is subjective, labor-intensive, and impractical for the prolonged multi-night monitoring required for clinical trials or home use. Recent studies using 2D and 3D infrared cameras have shown that automated analysis of REM sleep video can distinguish RBD from other sleep disorders with high accuracy,^11-14^ including a 2D camera study from our group achieving 91.9% accuracy.^13^

Here, we extend this approach to the first automated severity rating of RBD movements, as a step toward objective digital endpoints for symptomatic treatment trials and home monitoring.

## METHODS

### Subjects

Clinical video-polysomnography (vPSG) data from 214 patients (86 isolated RBD [iRBD], 17 secondary RBD, and 111 controls) collected at the Stanford Sleep Center between 2016 and 2022 were included in this study. Inclusion criteria included age ≥ 40 years old, evidence of REM sleep without atonia (RSWA) combined with a documented history of dream enactment behaviors, and absence of a diagnosed neurodegenerative disease or dementia.^12,15^ All diagnoses adhered to the International Classification of Sleep Disorders, third edition (ICSD-3).^16^ A total of 111 sex- and age-matched controls were randomly selected from the Stanford Sleep Center database. Data from secondary RBD patients (71.18 ± 7.82 years and 64.7% male) and controls (63.8 ± 9.2 years and 71.2% male) were used exclusively for pretraining the vision transformer on infrared movement patterns. Secondary RBD diagnoses included Parkinson’s disease (n=9), dementia with Lewy bodies (n=4), multiple system atrophy (n=2), multiple sclerosis (n=1) and chronic inflammatory demyelinating polyneuropathy (n=1). Data from control and secondary RBD participants were used exclusively to pretrain a deep-learning model as explained below. This study was approved by the Stanford University Institutional Review Board.

### Video-Polysomnography and Infrared 2D Camera Recordings

All recordings were acquired using the SomnoMedics (Randersacker, Germany) system following American Academy of Sleep Medicine (AASM) standards. The montage included electroencephalogram (EEG), electro-oculogram (EOG), and electromyogram (EMG) recordings of the mentalis, flexor digitorum superficialis, and anterior tibialis muscles, along with cardiac and respiratory channels (**Fig. 1**).^17^ Raw video was captured via a ceiling-mounted infrared two-dimensional (2D) infrared camera with a resolution of 704 × 576 pixels, a data rate of 100,000 kbps, and a frame rate of 30 frames per second (fps). The camera was positioned approximately 6 feet above and 3 feet away from the foot end of the bed in any of the 15 clinical rooms of the Stanford Sleep Center. Each recording was first scored in 30-second epochs by a sleep technologist, then reviewed and edited as needed by a board-certified sleep specialist. Periodic limb movements (PLMs) were identified using the SomnoMedics proprietary algorithm, while the RSWA index was manually derived from combined mentalis and flexor digitorum superficialis EMG activity in 30-second epochs according to SINBAR criteria.^18^

**Figure 1.**
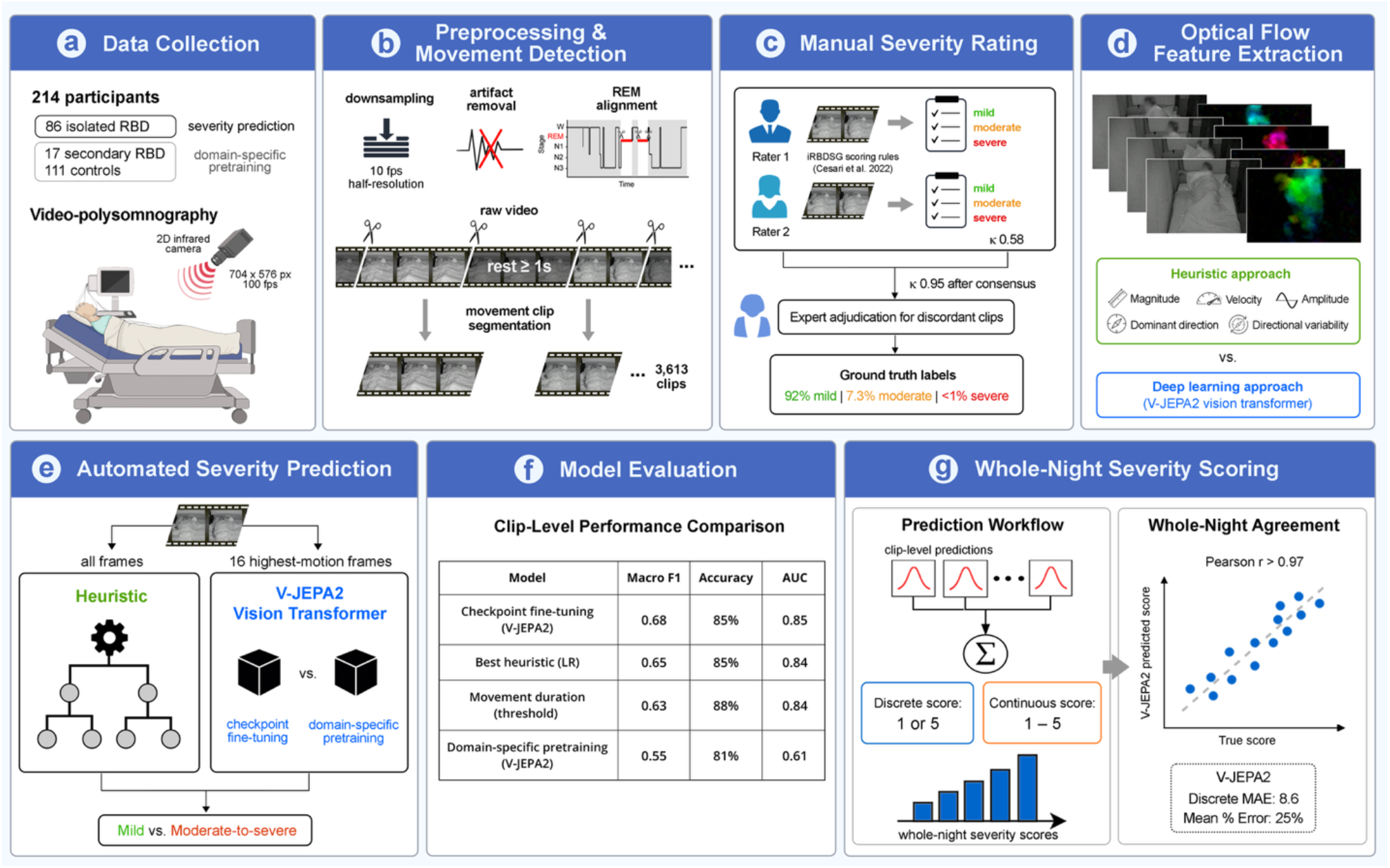
Overview of our study design and results. **a. Data collection**. Video-polysomnography with synchronized 2D infrared camera recordings from 214 participants: 86 isolated RBD patients for severity classification, 17 RBD patients and 111 controls for foundation model pretraining. **b. Preprocessing and movement detection**. Raw video was downsampled, optical-flow artifacts removed, and recordings aligned to REM sleep staging. Segmentation yielded 3,613 movement clips after exclusions. **c. Manual severity rating**. Two clinicians independently rated clips as mild, moderate, or severe per International RBD Study Group criteria, with consensus review and expert adjudication of unresolved cases. Inter-rater reliability improved from κ = 0.58 to κ = 0.95 after consensus. **d. Optical flow feature extraction**. Dense optical flow was computed per clip using DPFlow, yielding five heuristic features (magnitude, velocity, amplitude, dominant direction, directional variability) for the baseline classifier. **e. Automated severity prediction**. Two approaches were compared: a heuristic approach and V-JEPA2, a self-supervised video transformer, evaluated with checkpoint fine-tuning and domain-specific pretraining. Maximum optical-flow frame sampling was the optimal input strategy. **f. Model evaluation**. V-JEPA2 with checkpoint fine-tuning outperformed the best heuristic (logistic regression), movement duration, and domain-specific pretraining models in clip-level performance. **g. Whole-night severity scoring**. Clip-level predictions were aggregated into whole-night severity scores using discrete and continuous weighting schemes. Despite systematic overestimation, patient-level ordering was preserved (Pearson r > 0.97), with V-JEPA2 achieving a mean absolute error of 8.6 points and mean percentage error of 25%, compared to 20.0 points and 52% for the SVC.wsaeqw32

### Optical Flow and Data Preparation

Optical flow was utilized to quantify pixel-level displacement between consecutive frames. DPFlow was selected for its optimal balance between state-of-the-art accuracy and computational efficiency.^19,20^ For analysis, videos were downsampled to 10 fps and spatially resized to half-resolution, of their original width and height using OpenCV area-based interpolation. Several preprocessing steps were executed to ensure high-quality data input and minimize artifacts, noise, and irrelevant signals. First, file integrity was verified via checksums, and corrupted recordings were excluded from the experiments. All automatically flagged videos were manually reviewed to confirm their exclusion. Temporal smoothing using moving averages was applied to mitigate high-frequency noise. Principal component analysis (PCA) was subsequently performed on the pixel data to reduce computational load before optical flow extraction. Finally, we set a threshold (0–400) for acceptable movement ranges; frames exceeding these values, typically representing camera focus adjustments or light flickering, were flagged as outliers and removed to prevent skewed results in the downstream machine learning models. To isolate movements occurring during REM sleep, video recordings were aligned with vPSG staging data using temporal padding (**Supplementary Note 1**). Motion signals were smoothed after extraction by averaging optical flow magnitudes in 0.5 s bins, yielding one motion value per bin. To mitigate inter-individual variability arising from differences in camera setup, patient-level normalization was applied to each feature vector using baseline statistics derived from still segments, with z-score, IQR, and MAD scaling evaluated to account for the heavy-tailed and skewed distributions of the extracted features (**Supplementary Note 2)**.

### Segmentation of Movement Clips

Movement episodes were segmented according to protocols by Cesari et al.,^12^ defined by the transition from immobility to detectable motion and separated by at least one second of immobility. Dense optical flow was estimated between consecutive frames using a Sparse-to-Dense Optical Flow method.^21^ First, feature points were detected in each frame and tracked across subsequent frames to estimate sparse motion vectors. These sparse motion estimates were then interpolated to neighboring pixels to obtain a dense optical flow field. Frame-wise optical flow magnitudes were computed from the resulting dense flow and used to identify movement peaks and segment movement episodes within each clip.

### Manual Scoring for Movement Severity Labels

After processing, 4,099 movement clips were extracted during REM sleep. Two trained clinicians independently reviewed all clips to establish ground truth labels by rating movement severity as mild, moderate, or severe per IRBDSG guidelines.^9^ Movements with no risk of injury were classified as mild, intermediate movements as moderate, and forceful, vigorous, or violent movements capable of severely injuring the patient or bed partner (e.g., kicking or punching) as severe. Discordant ratings were re-reviewed jointly at a consensus meeting; clips that remained discordant were adjudicated by a senior expert. At each stage, clips for which REM sleep staging was uncertain were flagged and removed if a majority of reviewers deemed them ambiguous or clearly unsuitable for analysis.

### Class imbalance

The dataset exhibited marked class imbalance, with 92% of clips rated as mild. Moderate and severe movements were therefore merged into a single binary class (moderate-to-severe) to reduce class sparsity while preserving clinical relevance. Standard class-imbalance correction techniques were applied during model training, strictly within training folds to prevent information leakage.

### Movement Severity Modeling

Two distinct approaches for movement severity prediction were evaluated in this study: a heuristic model based on computed optical flow movement features and an approach utilizing V-JEPA2, a self-supervised video foundation model based on a Joint Embedding Predictive Architecture (JEPA) with a Vision Transformer (ViT) backbone.

### Heuristic modeling

#### Feature computation

Extending previous work on a diagnostic classifier, five clinically relevant heuristic movement features were derived from the optical flow of each extracted movement clip^13^: (1) movement magnitude, (2) velocity, (3) dominant motion direction, (4) directional variability, and (5) amplitude. Movement duration was also included as an additional feature. Features were summarized using robust statistics and aggregated into 0.5 s bins. Full mathematical definitions are provided in **Supplementary Note 3**.

#### Feature evaluation and selection

Strong correlation clusters were observed within several feature families, particularly those related to motion magnitude, exit fraction, angular displacement, and angular velocity (**Supplementary Fig. 1**). This was computed on the training and validation sets (3,023 clips from the 90/10 split) to avoid data leakage. Multicollinearity was reduced by removing one feature from each pair with a Pearson correlation coefficient greater than 0.9. This process identified 11 key features: clip duration; the mean and standard deviation of directional dispersion computed using circular variance; the mean, 80^th^ percentile, and 90^th^ percentile of motion velocity; the mean and standard deviation of dominant motion direction; and the mean and standard deviation of the fraction of pixels exhibiting motion above threshold.

#### Duration dominance and feature setup

Inspection of the resulting feature set revealed that clip duration exhibited substantially stronger class separability than all other retained features, motivating the definition of three complementary feature setups to disentangle its contribution. The first setup used duration alone as the simplest possible baseline. Given its observed distributional properties, this baseline was implemented as a single threshold classifier. In the ***easy***-split setting, the Macro F1 score was computed for every candidate threshold on each cross-validation fold, and the threshold maximizing the mean Macro F1 score across folds was selected and evaluated on the held-out test set. In the ***hard***-split setting, the same search was conducted over the combined training and validation sets, and the selected threshold was evaluated on the held-out test set. The second setup excluded duration entirely and retained only the remaining correlation-filtered features to assess whether they carried independent discriminative information. The third setup included all retained features, including duration, to quantify the extent to which its dominance degraded or masked the contribution of the remaining features.

#### Classifier training and hyperparameter optimization

Following feature selection, two complementary feature-processing strategies were investigated: one utilizing the retained features directly and another employing PCA to maintain a fixed proportion of explained variance. PCA was fit exclusively on training data to prevent information leakage, with feature contributions quantified as weighted absolute loadings to ensure interpretability of the reduced feature space. Five classical supervised learning models, including LR, SVC, Random Forest, AdaBoost, and XGBoost, were evaluated. All models were trained on standardized feature vectors using pipeline-based implementations to ensure consistent preprocessing. For each algorithm, a dedicated hyperparameter grid was defined, covering regularization strength and penalty type for LR; kernel choice and margin parameters for SVC; tree depth and ensemble size for Random Forest and AdaBoost; and learning rate and boosting depth for XGBoost.

### Foundation model

Several foundation model architectures were evaluated for feature extraction and visualization, including DINOv3,^22^ before V-JEPA2, a self-supervised video foundation model based on a Joint Embedding Predictive Architecture (JEPA) with a Vision Transformer (ViT) backbone was selected for its superior ability to model temporal dynamics and robustness to static-noise rejection in infrared video.^23^ Severity classification was explored through two approaches: fine-tuning a publicly released pretrained checkpoint and domain-specific self-supervised pretraining from scratch.

#### Checkpoint-based fine-tuning

The first set of experiments used V-JEPA2 initialized from the publicly released checkpoint “vjepa2-vitl-fpc16-256-ssv2 (https://huggingface.co/facebook/vjepa2-vitl-fpc16-256-ssv2)”. A baseline was established using a naïve sampling strategy with a frozen encoder, in which only the classification head was fine-tuned. Each movement clip was represented by 16 uniformly selected frames and processed in a batch size of 16. To mitigate severe class imbalance, weighted cross-entropy loss with inverse-frequency class weights was applied. Additional sampling refinements included merging moderate and severe classes into a single category. Given the substantial domain mismatch between natural video pretraining datasets and infrared sleep recordings, the entire V-JEPA2 model was subsequently unfrozen and fully fine-tuned. Overfitting was controlled through weight decay, early stopping, and a learning-rate warm-up period comprising 10% of total steps, with model selection based on Macro F1 score or AUC. Hyperparameters were optimized via a 3-fold cross-validation grid search.

A major challenge in this task was selecting a fixed 16-frame input from movement clips of variable duration. In addition to the baseline naïve sampling, four distinct frame-sampling strategies were evaluated to determine the optimal leverage of temporal information and motion cues. Uniform sampling partitioned each clip into 16 equally spaced time points, with one representative frame selected from each time point to ensure coverage of the full clip. Peak-flow window (uniform) identified a 3-second window with the highest average optical flow magnitude and sampled 16 frames at equal time intervals within that period. Peak-flow window (random) utilized the same high-motion window but randomly sampled 16 frames randomly within it. Finally, the maximum optical flow strategy selected the 16 frames exhibiting the highest optical flow magnitudes relative to their preceding frames (**Supplementary Fig. 2**). This approach explicitly targeted peak-motion events without any predefined temporal windows. Frame-level magnitudes for this selection were initially estimated using the Farnebäck dense method,^24^ which showed high consistency with DPFlow results, matching over 80% of identified peak-motion frames.

### Domain-specific pretraining

#### Movement extraction for pretraining

To support vision-transformer pretraining on infrared movement patterns, whole-night data (including wake, non-REM and REM sleep) from all cohorts (86 iRBD, 17 secondary RBD cases, and 111 controls) were used, yielding more than 70,000 movement clips. Patient-adaptive movement thresholds between the 93rd and 97th percentiles of the smoothed magnitude distribution were applied to account for inter-individual variability in movement activity, with stricter thresholds for low-activity sleepers to minimize false positives (**Supplementary Note 4**).

#### Rationale for domain-specific pretraining

The specialized nature of the dataset in this study (e.g., low-light, grayscale infrared recordings with subtle motion and background noise) differed substantially from the large-scale natural video datasets used for V-JEPA2 pretraining, which consisted entirely of RGB internet video with no infrared content across approximately 22 million clips. The performance improvement observed after unfreezing the encoder during fine-tuning suggested a fundamental distribution mismatch between the original pretrained representations and the target domain. This prompted an investigation into custom self-supervised pretraining of V-JEPA2 from scratch to determine whether establishing domain-specific representations could further enhance downstream movement severity classification.

#### Pretraining procedure

For pretraining, movement clips were extracted using a lower optical flow threshold than that used for classification to capture subtle motions and increase the diversity of the self-supervised learning data. Pretraining adhered to the two-stage framework proposed in the original V-JEPA2 architecture. The first stage utilized 16 frames per clip to establish basic temporal representations, whereas the second stage employed 32 frames per clip and an extended training duration to capture more complex temporal dependencies. Initial replication of the original training duration and hyperparameters resulted in a first-stage trend that exhibited an initial decrease followed by a gradual increase in expected loss, a behavior consistent with the custom self-supervised objective (**Supplementary Fig. 3a**). However, the second stage was marked by rapid convergence and unstable loss oscillations, suggesting insufficient convergence before increasing temporal complexity (**Supplementary Fig. 3b**). To rectify this, the duration of the first stage was extended to improve convergence on simpler temporal patterns before progression to the second stage (**Supplementary Fig. 4a**). This scaled schedule achieved a more stable decrease in loss and improved overall convergence, validating the effectiveness of the modified training procedure (**Supplementary Fig. 4b**).

#### Fine-tuning for severity classification

Following pretraining, an attention-based classification head identical to that used in the original V-JEPA2 checkpoint was integrated for fine-tuning for movement severity classification. Three fine-tuning strategies were systematically evaluated: maintaining a frozen encoder, fully unfreezing the encoder, and a two-stage fine-tuning approach in which the classification head was first trained with a frozen encoder prior to joint optimization of both components. Results from the two-stage fine-tuning strategy utilizing uniform frame sampling were included as a primary reference for model comparison.

#### Data splitting

Hyperparameter optimization and model evaluation were performed using two stratified data-splitting strategies: an easy split and a hard split.

In the easy split, stratification was performed at the clip level, preserving class proportions across training and test sets. Clips from the same patient could appear in different subsets, maximizing the available training data but potentially producing optimistic performance estimates due to within-patient information overlap. Hyperparameters were optimized on the training data using GridSearchCV with stratified 5-fold cross-validation. Final performance was assessed using 5-fold cross-validation, with five folds of 18% each, and an additional independent 10% held-out test set.

In the hard split, partitioning was performed at the patient level to prevent information leakage. All clips from a given patient were assigned to a single subset, ensuring that validation and test data came from entirely unseen patients. Because of substantial class imbalance and unequal numbers of clips per patient, a single stratified 80/10/10 training/validation/test split was used; a 5-fold cross-validation scheme with an additional independent 10% test set was not feasible while preserving class proportions.

#### Continuous severity rating and whole-night severity scores

Clip-level severity predictions for REM sleep movements were aggregated into a single scalar score to summarize whole-night severity for each patient. In the discrete severity setting, predicted labels were assigned fixed point values: 1 point for mild movements and 5 points for moderate-to-severe movements. In the continuous severity setting, clip-level predicted probabilities were converted into per-clip point contributions before aggregation, using model-specific scaling functions to preserve comparability with the discrete scoring scheme (**Supplementary Note 5**).

Whole-night severity scores were reported for the 14 patients included in the hard-split validation and test sets. This analysis was not performed in the easy-split setting because night-level scoring required all clips from a given patient to remain within the same partition. Night-level performance was summarized using absolute error, percent error, bias, overestimation and underestimation rates, and per-clip normalized error metrics (**Supplementary Note 6**).

#### Statistical analysis

Normality of the continuous variables was assessed using the Shapiro–Wilk test and Q-Q plots. Group comparisons were performed using independent t-tests or Mann–Whitney U tests, as appropriate. Categorical variables were analyzed using χ2 or Fisher’s exact tests. A two-sided p value < 0.05 was considered statistically significant. Inter-rater reliability for movement severity ratings across clips was evaluated using percent agreement and unweighted Cohen’s kappa (κ), both before and after consensus review. Cohen’s κ was also reported as a complementary chance-corrected measure of model-consensus agreement for predicted versus consensus ground-truth labels. All statistical analyses were conducted using R version 4.5.2 (R Core Team, 2025).

## RESULTS

### Study population

Characteristics of the iRBD cohort are presented in **Table 1** (see **Supplementary Table 1** for all cohorts).

**Table 1.** Demographic, clinical, and polysomnographic features of the iRBD cohort (n = 86). AHI3A, apnea-hypopnea index scored using hypopneas associated with ≥3% oxygen desaturation or arousal; NREM, non-rapid eye movement; OSA, obstructive sleep apnea; PAP, positive airway pressure; PLM, periodic limb movement; REM, rapid eye movement; RLS, restless legs syndrome; RSWA, REM sleep without atonia; SD, standard deviation.

|  |  |
| --- | --- |
| **Demographics** |  |
| Age, mean (SD) | 65.0 (8.6) |
| Male, n (%) | 68 (79.1) |
| **Sleep disorders** |  |
| Insomnia, n (%) | 4 (4.7) |
| RLS, n (%) | 11 (12.8) |
| Sleepwalking, n (%) | 4 (4.7) |
| Narcolepsy type 1, n (%) | 0 (0) |
| AHI3A $\geq 15$, n (%) | 21 (24.4) |
| PAP therapy, n (%) | 25 (40.3) |
| **Sleep variables** |  |
| Total sleep time, min, mean (SD) | 381.4 (78.8) |
| NREM sleep time, min, mean (SD) | 309.5 (70.0) |
| REM sleep time, min, mean (SD) | 72.2 (43.7) |
| AHI3A, mean (SD) | 11.8 (10.9) |
| REM AHI, mean (SD) | 10.8 (10.8) |
| PLM index, mean (SD) | 30.7 (36.2) |
| PLM index $\geq 15$, n (%) | 48 (55.8) |
| RSWA index, mean (SD) | 0.64 (0.26) |

### Movement severity distribution and inter-rater reliability

Of the 4,099 movement clips, 486 were excluded upon manual review — due to artifacts (e.g., clinic personnel crossing the room) or clinical grounds that movement occurred during wakefulness (e.g., slow repositioning, scratching) — yielding 3,613 retained clips. Of these, 3,329 (92.1%) were rated as mild, 265 (7.3%) as moderate, and 19 (0.6%) as severe (**Supplementary Fig. 5**). The mean whole-night severity score was 68.8 ± 82.9 points (**Supplementary Fig. 6**).

Initial blinded review yielded 85.1% agreement (κ = 0.58). Among the 612 discordant clips, 412 (67.3%) reflected mild-moderate disagreement, 24 (3.9%) moderate-severe disagreement, and 176 (28.8%) uncertainty about REM sleep staging. Following consensus review, agreement improved to 98.5% (κ = 0.95), with 61 clips (1.5%) requiring senior expert adjudication.

### Heuristic model

Among all motion-derived features, clip duration had the highest permutation importance, followed by mean angular variance, while the remaining features clustered around or below zero (**Fig. 2a**). When duration was excluded, importance shifted toward movement amplitude mean and standard deviation, mean motion angle standard deviation, and upper-tail velocity (**Fig. 2b**).

**Figure 2.**
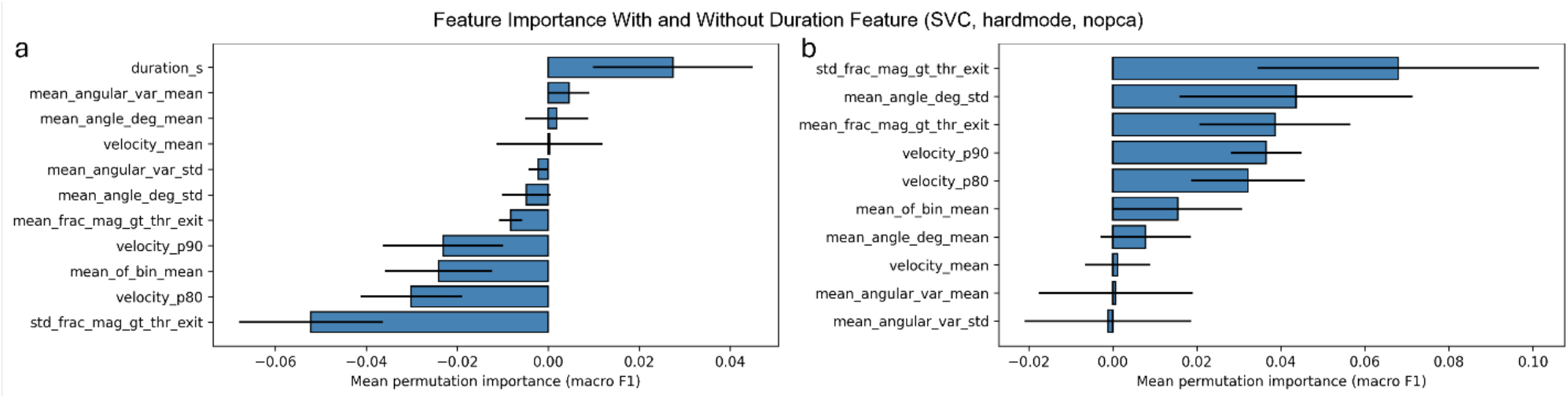
Permutation feature importance for the Support Vector Classifier (SVC) in the hard-split setting without principal component analysis (PCA), shown for (a) the model including and (b) excluding duration. Permutation feature importance, scored using Macro F1, measures the drop in performance after a random shuffling of feature values while leaving the trained model unchanged. Positive values indicate features on which the model relies on, near-zero values indicate uninformative features, and negative values indicate features that act as noise.

The duration-only threshold classifier performed moderately in the easy split, achieving a Macro F1 of 0.69 and AUC of 0.78 (**Table 2; Supplementary Tables 2 and 3**). Recall and precision for detecting moderate-to-severe movements were 46% and 42%, respectively. Macro-averaged across all classes, these reached 70% and 69%. Performance was modestly lower in the hard split, with a Macro F1 of 0.63 and AUC of 0.84; recall and precision for moderate-to-severe detection were 34% and 32%, and macro-averaged values were 64% and 63%. Feature-based models not including movement duration performed similarly with best Macro F1 scores and AUC of 0.56 and 0.74 in the easy split (RF), and 0.65 and 0.83 in the hard split (SVC). Combining all features and movement duration hardly affected performance of either approach alone.

**Table 2.**
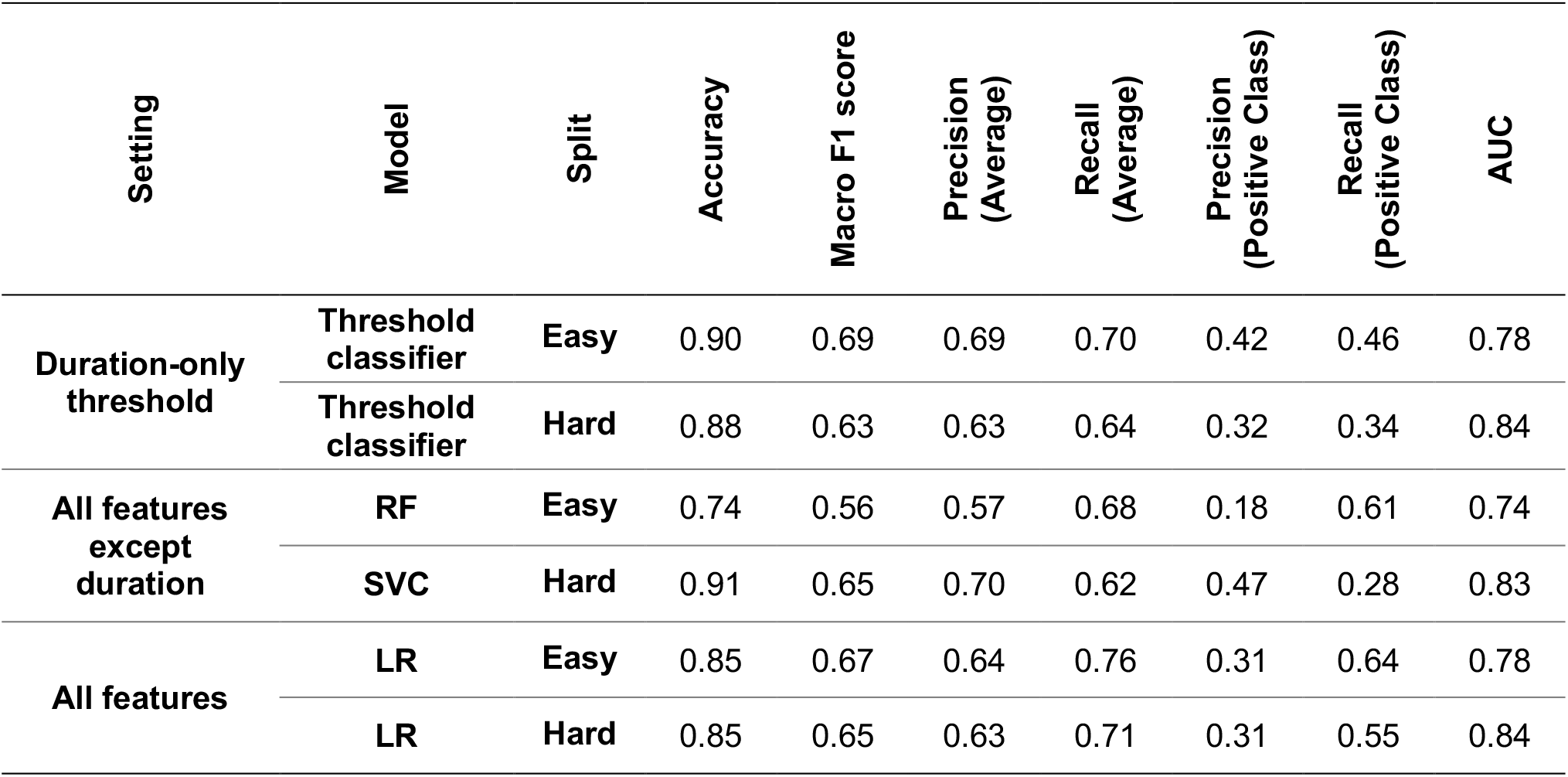
Heuristic analyses across duration-only and feature-based models. Results are shown for the easy and hard split settings and reflect the best-performing model and preprocessing configuration for each strategy (full results across all configurations reported in Supplementary Tables 2 and 3). All models used raw optical-flow features without normalization or principal component analysis. Positive class is moderate-to-severe movements. The duration-only baseline applied a single threshold to untransformed clip duration. Feature-based analyses used all retained features, with and without duration. AUC, area under the curve. LR, Logistic Regression. RF, Random Forest. SVC, Support Vector Classifier.

### Foundation model

For both checkpoint-based and domain-specific V-JEPA2,^23^ best performances were achieved using the maximum optical flow-based sampling strategy, slightly outperforming any other sampling method (**Supplementary Tables 4 and 5**). Checkpoint-based fine-tuning achieved the highest Macro F1 score and accuracy of 0.76 and 93% in the easy split setting, and 0.68 and 85% in the hard split setting (**Table 3 and Fig. 3**). In comparison, domain-specific two-stage fine-tuning achieved a Macro F1 score and accuracy of 0.73 and 90%, and 0.55 and 81% in the easy and hard split settings, respectively. Detailed results of frozen and unfrozen encoder strategies yielded Macro F1 scores of 0.49 and 0.69, respectively.

**Table 3.** Performance of the two best-performing V-JEPA2 approaches. Both models use maximum optical flow-based frame sampling. AUC, area under the curve.

| Model | Split | Accuracy | Macro F1 | Precision | Recall | AUC |
| --- | --- | --- | --- | --- | --- | --- |
| Checkpoint-based | Easy | 0.93 | 0.76 | 0.75 | 0.77 | 0.90 |
|  | Hard | 0.85 | 0.68 | 0.65 | 0.75 | 0.85 |
| Domain-specific with Two-stage fine tuning | Easy | 0.90 | 0.73 | 0.69 | 0.80 | 0.86 |
|  | Hard | 0.81 | 0.55 | 0.55 | 0.57 | 0.61 |

**Figure 3.**
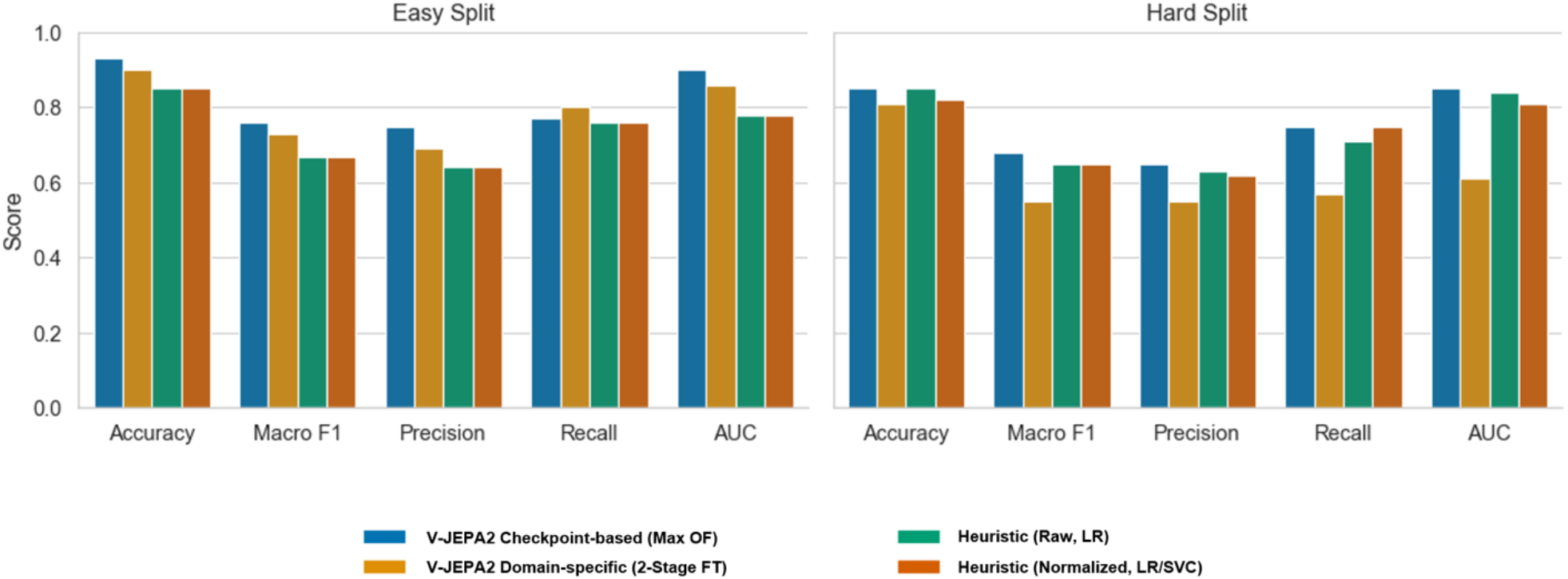
Comparisons of best performing foundation and heuristic models. AUC, area under the curve; FT, fine tuning; LR, Logistic Regression; OF, optical flow; SVC, Support Vector Classifier.

In the two-class hard-split test set, model-consensus agreement was moderate, with a Cohen’s κ of 0.42 between foundation model predictions and consensus ground-truth labels. Because this estimate was based on approximately 300 clips and used post-consensus labels as ground truth, it should be interpreted as complementary to Macro F1 rather than as a standalone performance measure.

### Continuous severity rating and whole-night severity scores

At the clip level, both foundation and SVC models exhibited positive bias (**Table 4**). For SVC models, the mean absolute error (MAE) was 0.60 for discrete ratings and 0.77 for continuous ratings. For foundation models, validation-set MAEs were 0.31 for discrete ratings and 0.90 for continuous ratings.

**Table 4.** Clip-level performance comparison of vision-based and SVC models using discrete and continuous ratings. Errors are reported as points per clip. All confidence intervals are 95% patient-level bootstrap confidence intervals (n = 7 nights per split). The V-JEPA2 validation set and SVC test set contained video clips from the same subjects. AE, absolute error; SVC, Support Vector Classifier.

| Model | Ratings | Mean AE | Median AE | Bias |
| --- | --- | --- | --- | --- |
| **V-JEPA2**<br>(Hard split,<br>validation set) | **Discrete** | 0.31 [0.07, 0.61] | 0.03 [0.00, 0.71] | +0.31 [0.05, 0.61] |
|  | **Continuous** | 0.90 [0.61, 1.23] | 0.68 [0.56, 1.44] | +0.90 [0.61, 1.23] |
| **V-JEPA2**<br>(Hard split,<br>test set) | **Discrete** | 0.36 [0.15, 0.63] | 0.23 [0.13, 0.57] | +0.36 [0.15, 0.63] |
|  | **Continuous** | 0.88 [0.54, 1.23] | 0.91 [0.48, 1.13] | +0.88 [0.54, 1.23] |
| **SVC**<br>(Hard split,<br>test set) | **Discrete** | 0.60 [0.30, 0.97] | 0.50 [0.16, 0.72] | +0.60 [0.30, 0.97] |
|  | **Continuous** | 0.77 [0.52, 1.09] | 0.73 [0.37, 0.90] | +0.77 [0.52, 1.09] |

At the night level, the MAEs for SVC models were 20.0 for discrete ratings and 28.9 for continuous ratings (**Table 5**). Corresponding MAEs for foundation models were 8.6 and 31.7, respectively. Mean percentage errors (MPEs) for SVC models were 52% for discrete ratings and 65% for continuous ratings, whereas MPEs for foundation models were 25% and 72%, respectively.

**Table 5.** Night-level error metrics computed across nights. Absolute errors are reported as total points per night, while percentage errors reflect relative deviation from the true night score. All confidence intervals are 95% patient-level bootstrap confidence intervals. The V-JEPA2 validation set and SVC test set contained video clips from the same subjects. AE, absolute error; SVC, Support Vector Classifier.

| Model | Ratings | Mean AE | Median AE | Bias | Mean % error | Median % error |
| --- | --- | --- | --- | --- | --- | --- |
| **V-JEPA2**<br>(Hard split,<br>validation set) | **Discrete** | 8.6<br>[1.7, 18.3] | 4.0<br>[0.0, 12.0] | +7.4<br>[-0.6, 17.7] | 25%<br>[5%, 49%] | 2%<br>[0%, 67%] |
|  | **Continuous** | 31.7<br>[16.7, 49.3] | 22.5<br>[12.6, 61.8] | +31.7<br>[16.7, 49.3] | 72%<br>[45%, 108%] | 63%<br>[38%, 96%] |
| **V-JEPA2**<br>(Hard split,<br>test set) | **Discrete** | 14.3<br>[5.1, 28.0] | 8.0<br>[4.0, 16.0] | +14.3<br>[5.1, 28.0] | 28%<br>[12%, 47%] | 20%<br>[11%, 57%] |
|  | **Continuous** | 42.6<br>[14.2, 87.8] | 26.1<br>[14.3, 37.3] | +42.6<br>[14.2, 87.8] | 70%<br>[44%, 94%] | 67%<br>[38%, 113%] |
| **SVC**<br>(Hard split,<br>test set) | **Discrete** | 20.0<br>[9.7, 33.2] | 16.0<br>[4.0, 28.0] | +20.0<br>[9.7, 33.2] | 52%<br>[23%, 93%] | 33%<br>[10%, 60%] |
|  | **Continuous** | 28.9<br>[14.3, 46.1] | 25.6<br>[8.6, 49.5] | +28.9<br>[14.3, 46.1] | 65%<br>[37%, 100%] | 55%<br>[26%, 73%] |

Pearson’s correlations between predicted and ground-truth whole-night severity scores across discrete and continuous rating settings are shown in **Fig. 4**.

**Figure 4.**
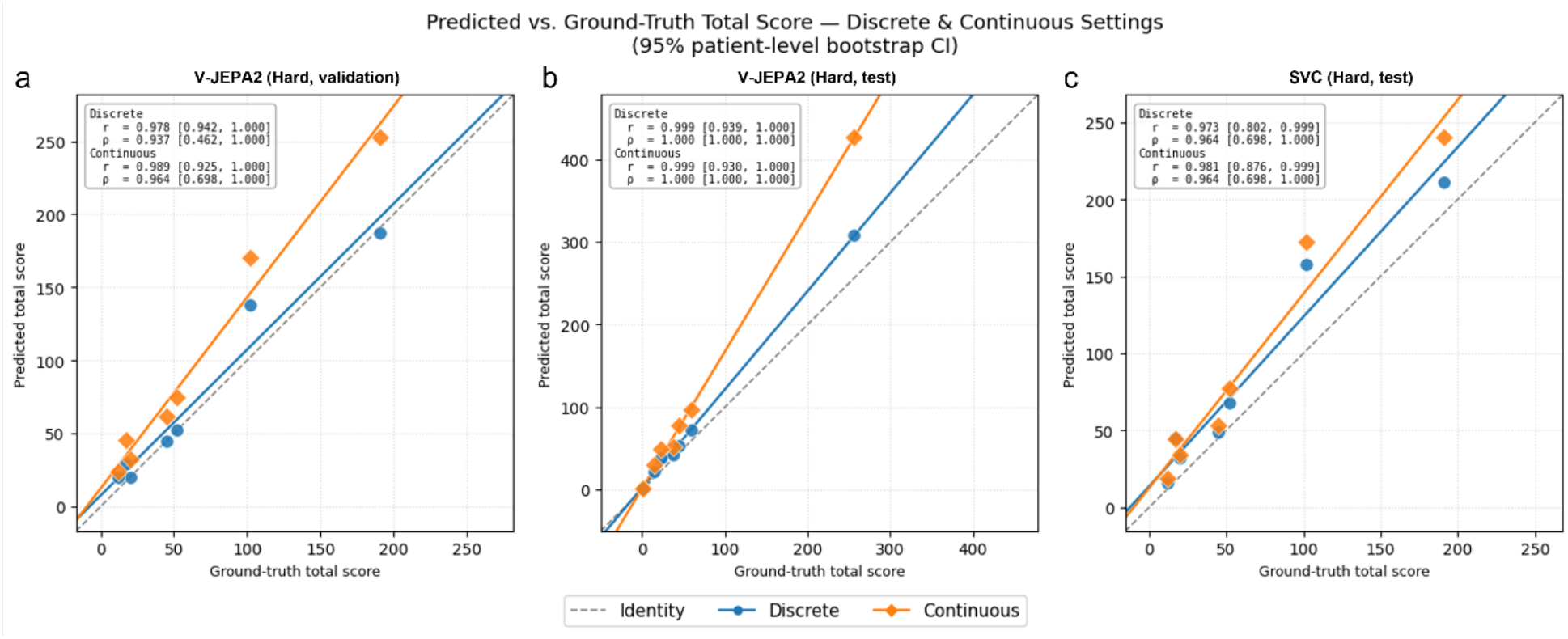
Correlation between predicted and ground-truth whole-night severity scores across discrete and continuous rating settings. Results are shown for the V-JEPA2 validation set, V-JEPA2 test set, and SVC test set (n = 7 nights per split). Pearson r and Spearman ρ values are reported with 95% patient-level bootstrap confidence intervals (CIs). The V-JEPA2 validation set and SVC test set contain video clips from the same subjects. SVC, Support Vector Classifier.

## DISCUSSION

In this study, we developed and evaluated two complementary computer vision approaches for automated severity rating of RBD movements using infrared video recorded during REM sleep in a laboratory setting. Several findings emerged from this exercise. First, movement severity could be predicted from video with clinically meaningful performance despite substantial heterogeneity in sleep laboratory recordings and the strong class imbalance inherent to RBD severity data. Second, a vision-based approach outperformed heuristic optical flow-derived models at the clip level, particularly when frame selection was focused on peak motion. Third, although automated clip-level classification showed promising performance, translation of these predictions into continuous and whole-night severity scores was less stable, indicating that night-level aggregation is feasible but not yet sufficiently reliable for use as a standalone endpoint.

These findings are significant because the field currently lacks efficient objective outcome measures for RBD treatment trials. This remains a major barrier to therapeutic development, because such tools could transform clinical care by enabling more precise treatment titration, especially for patients who live or sleep alone and therefore cannot rely on bedpartner observations. Indeed, assessment of symptom severity now depends on patient and bedpartner reports, despite well known susceptibility to recall bias, expectancy effects, and marked night-to-night variability. Although analysis of REM sleep video data provides an objective measure of symptom severity, manual scoring is labor-intensive and impractical for prolonged monitoring, particularly in home settings. Our findings support the feasibility of automating videoprocessing, as computer vision would operationalize video-based severity assessment.

A strength of the present work is that it addresses both interpretability and performance. Among the optical flow-derived heuristic measures, clip duration was the dominant predictor of severity, likely because duration reflects persistence of supra-threshold motion and therefore functions as a cumulative motion measure. However, duration-independent motion descriptors, such as velocity, directional dispersion, and the spatial extent of movement, retained discriminative value when analyzed separately, suggesting that severity is also encoded in motion structure. Additionally, the relatively superior performance of V-JEPA2 with high-motion frame selection suggests that deep video representations can capture more complex patterns, particularly when attention is focused on the most informative motion epochs rather than averaged across the full clip. Moreover, learned video representations likely encode richer temporal structure, body configuration changes, and spatiotemporal patterns that are difficult to summarize using a limited set of engineered features. This may explain why maximum optical flow frame sampling performed best.

The performance difference between the easy and hard splits is also informative. As expected, allowing clips from the same patient to appear across dataset partitions yielded more favorable estimates, whereas patient-level partitioning provided a more stringent test of generalization. Performance remained largely preserved in the hard split, particularly for the foundation model, indicating that the learned representations were not driven solely by patient-specific idiosyncrasies. Notably, V-JEPA2’s performance advantage over the best heuristic model (LR) was larger in the easy split (0.76 vs. 0.67) than in the hard split (0.68 vs. 0.65), although its Macro F1 declined more steeply across splits (–0.08 vs. –0.02). This suggests that part of the foundation model’s easy-split advantage reflects within-patient regularities rather than generalizable severity representations. The modest 0.03 Macro F1 margin in the hard split underscores the importance of external validation across centers and recording conditions before conclusions can be drawn.

The finding that maximum optical flow-frame sampling achieved the best performance is conceptually appealing (**Table 3 and Fig. 4**). Severity ratings in RBD are generally driven by the most forceful or potentially injurious segments of a dream-enactment episode rather than by its average content over time. By preferentially selecting frames around peak motion, the model appears to capture the most discriminative portion of the clip, likely explaining its advantage over uniform and window-based sampling strategies and supporting the clinical plausibility of the approach. More broadly, this finding suggests that temporal sampling is an important design parameter in sleep video analysis and can substantially influence classification performance.

Our results also highlight the importance of adapting video models to the sleep-laboratory setting. While fine-tuning a pretrained general video model yielded strong performance, performance improved further when the full model was fine-tuned and when additional pretraining was performed on infrared sleep recordings. This suggests that features learned from natural and general-purpose video do not fully transfer to sleep video, which differs substantially in both appearance and motion characteristics. Infrared sleep recordings are grayscale, low-light, visually repetitive, and characterized by subtle movements against static backgrounds. Despite being trained on only 70,000 clips compared to the 22 million clips used for V-JEPA2 pretraining, the domain-specific model achieved a Macro F1 of 0.73 in the easy split — only 0.03 below checkpoint fine-tuning. This suggests that domain relevance can partially compensate for data volume, and that a larger infrared sleep video dataset could potentially close this gap further or even surpass the checkpoint-based approach. More broadly, these findings suggest that foundation models provide a useful starting point for sleep medicine applications, but robust performance may still depend on domain-specific pretraining and carefully curated datasets.

The inter-rater reliability data provide essential context for interpreting both the ground truth on which this study rests and the performance levels the automated models achieved. Before consensus, two independent expert clinicians agreed on only 58% of cases beyond chance (κ = 0.58), revealing meaningful disagreement concentrated precisely at the boundary between mild and more severe movements — the exact distinction this study set out to automate. This reflects a fundamental ambiguity in the current IRBDSG framework, which relies on visual judgment without quantitative anchors. Reaching κ = 0.95 required a structured consensus meeting and expert adjudication of 61 residual clips, a process that is not scalable to the clip volumes required for longitudinal monitoring or multi-site trials. These observations carry two important implications. First, model performance should be benchmarked against pre-consensus human agreement rather than a hypothetical perfect scorer: a Macro F1 of 0.76 operates at the boundary of inter-human variability, and some apparent model errors at the mild-to-moderate boundary may reflect genuine perceptual ambiguity rather than model failure. Second, automated scoring offers an advantage beyond speed: by applying consistent criteria uniformly across every clip, it eliminates the rater fatigue and criterion drift that accompany large-scale manual annotation. The moderate initial inter-rater agreement is therefore not only a limitation to acknowledge but a scientific justification for the approach, underscoring why manual scoring cannot simply be scaled up.

This study also explored continuous severity rating using an automated algorithm. While whole-night severity scores derived from the continuous ratings showed higher MAEs than discrete ratings, correlations between predicted and ground-truth scores remained high across models and scoring formulations, with Pearson correlation coefficients exceeding 0.97 (**Fig. 3**). However, the night-level analyses warrant cautious interpretation. While discrete whole-night severity scores were more accurate than continuous scores, especially for the foundation model, percentage errors remained substantial. This likely reflects error propagation across clips and imperfect probability calibration. Thus, our study establishes proof of concept for automated night-level scoring, while also demonstrating that improved calibration and aggregation strategies will be necessary before such scores can serve as robust endpoints for clinical trials. In particular, models optimized for clip-level Macro F1 score may not be optimal for patient-level or night-level severity estimation. Future work should explicitly optimize for night-level agreement, treatment responsiveness, and test-retest reliability.

Several limitations should be acknowledged. First, this was a single-center study based on infrared 2D sleep laboratory recordings acquired under non-uniform video conditions over several years. Although this heterogeneity may improve ecological robustness, it also introduces variability related to camera angle, focus, and lighting conditions. Second, synchronization between video and polysomnography was imperfect in a substantial portion of recordings, necessitating pragmatic use of a padded REM set for the primary analysis. Despite exclusion of approximately 12% of clips as being unlikely to be from REM sleep, it may have introduced some contamination from non-REM or wake movements and thereby reduced signal specificity. Third, the iRBD REM movement clips used for supervised fine-tuning in the domain-specific pretraining approach were also included in the self-supervised pretraining stage. Although no severity labels were available during pretraining, the encoder was exposed to the visual content of these clips prior to fine-tuning, which may have facilitated learning of patient-specific representations and inflated performance estimates for this approach relative to a fully held-out evaluation. Fourth, the class distribution was highly skewed, with more than 92% of clips rated as mild and <1% as severe. This limited the ability to model the full severity spectrum and necessitated a binary reformulation. Last, the ground truth itself was based on expert visual scoring rather than on an external physiologic or injury-based criterion; accordingly, the model learned expert ratings, not necessarily the underlying biological severity of RBD.

These limitations are balanced by several strengths. The study included a relatively large number of manually reviewed REM movement clips, rigorous consensus labeling, explicit attention to severity information leakage through both easy and hard splits, and a head-to-head comparison between interpretable feature-based methods and modern self-supervised video models. The incorporation of matched controls for pretraining is also a strength, as it exposed the vision transformer to a broader distribution of infrared sleep movements than would have been available from labeled RBD clips alone.

The present work represents a meaningful step toward objective, scalable digital endpoints for RBD. A fully deployable system would require reliable REM sleep identification, calibrated severity probabilities, stable night-level aggregation, and longitudinal tracking across repeated home recordings — capabilities that future studies should systematically validate. Beyond clinical trials, such tools could help phenotype patients more precisely and provide a granular bridge between symptom burden and underlying synucleinopathy progression.

In conclusion, automated severity rating of RBD movements from infrared video is feasible and performs best with vision-based spatiotemporal modeling. Further work is needed to improve calibration, preserve ordinal severity structure, and strengthen night-level scoring before this approach can serve as a validated endpoint for clinical trials or home monitoring.

## Acknowledgements

Open access funding was provided by the Mount Sinai Department of Neurology (IS130501152). The funding sources had no involvement in the study design, in the data extraction, in the writing, or in the decision-making.

## Author Contributions

J.H. and N.P. wrote the first draft of the manuscript. J.H. and B.R. reviewed and rated the severity of movement on video clips. N.P., A.G., and A.B. contributed to the acquisition and analysis of data, as well as preparing figures. K.R. edited the manuscript and created figures. A.S. reviewed and rated the severity of movement on video clips that had initially discordant ratings and reviewed the manuscript. S.M., O.S-P., and E.M. contributed to the acquisition of data. M.A., A.B-K., M.C., A.S., A.A., and E.D. contributed to data acquisition, conception and design of the study and performed the final review and editing of the manuscript. All authors read and approved the final paper.

## Data Availability

The data that support the findings of this study are available on request from the corresponding author. The raw video data is not available due to containing information that could compromise the privacy of research participants.

## Code Availability

The analysis code used in this study is available from the corresponding author upon reasonable request. It is not publicly available because it is project-specific and depends on non-public raw video inputs.

