## Supplementary Materials for "Vision-Based Automated Severity Rating of REM Sleep Behavior Disorder: From Heuristic Features to Foundation Models"

**Supplementary Note 1: Video-vPSG synchronization**

To restrict analyses to movements occurring during REM sleep, video timestamps were aligned with sleep-stage annotations using start-time alignment (the temporal padding), assuming simultaneous initiation of video and sensor channel recordings. Visual inspection suggested discrepant start times in roughly 50% of the samples. Exploratively, lights-off alignment, matching the vPSG "lights-off" timestamp to the corresponding video event, was performed to maximize the number of movements within the possible REM stage.

To account for this limitation in synchronization, REM stage intervals were padded by a maximum plausible mismatch capped at two minutes, generating a "low-confidence" REM movement set. A secondary "high-confidence" set utilized strict priority-based staging without padding; however, under these criteria, approximately 40% of labeled movements fell outside REM sleep. Therefore, the low-confidence set was used for the primary analyses to maintain consistency with the original model evaluation.

Using video-vPSG synchronization with temporal padding and the low-confidence REM movement set, 4,099 REM-stage movement clips were generated in this study. The full dataset used for V-JEPA 2 pretraining did not require synchronization, as clips were extracted across all sleep stages to maximize training-signal diversity.

**Supplementary Note 2: Denoising, Smoothing, Normalization**

**Denoising and smoothing**

To address the inherently higher background noise in infrared video relative to the standard red, green, and blue (RGB) video, several video denoising approaches were evaluated, including bilateral filtering,^1^ non-local means filtering,^2^ and Gaussian filtering^3^ in both grayscale and RGB formats. Paired t-tests comparing the average optical flow magnitude metrics before and after denoising showed no significant reduction in noise metrics. While grayscale bilateral filtering improved visual clarity without sacrificing the optical flow magnitudes, denoising was not applied before extracting flows.

**Patient-level normalization**

To mitigate inter-individual variability arising from differences in camera angles, magnification, and background configuration, several patient-level normalization methods were explored, including z-score, interquartile range (IQR), and median absolute deviation (MAD) scaling. These approaches were evaluated because the extracted features exhibited heavy-tailed and skewed distributions that violated the normality assumptions underlying standard z-score normalization. Baseline statistics from still segments were computed for each patient to standardize feature vectors and reduce specific background noise, including the baseline mean (𝑏_𝜇_), baseline standard deviation (𝑏_𝜎_), baseline median (𝑏_𝑚_), baseline interquartile range (𝑏_IQR_), and baseline median absolute deviation (𝑏_MAD_). A small constant 𝜀 > 0 was used to avoid division by zero.

Magnitude features

- Raw: $x'=x$
- m0 (median-centered): $x'=x-b_{m}$
- IQR (robust scale, centered): $x'=\frac{x-b_{m}}{b_{\mathrm{IQR}}+\varepsilon}$ (robust scale only for standard deviation features)
- MAD (robust scale, centered): $x'=\frac{x-b_{m}}{b_{\mathrm{MAD}}+\varepsilon}$ (robust scale only for standard deviation features)

Velocity features

- Raw: $x'=x$
- IQR (robust scale only): $x'=\frac{x}{b_{\mathrm{IQR}}+\varepsilon}$
- MAD (robust scale only): $x'=\frac{x}{b_{\mathrm{MAD}}+\varepsilon}$

Amplitude features

- Raw: $x'=x$
- z-score (robust scale, centered): $x'=\frac{x-b_{\mu}}{b_{\sigma}+\varepsilon}$ (robust scale only for standard deviation features)
- IQR (robust scale, centered): $x'=\frac{x-b_{m}}{b_{\mathrm{IQR}}+\varepsilon}$ (robust scale only for standard deviation features)
- MAD (robust scale, centered): $x'=\frac{x-b_{m}}{b_{\mathrm{MAD}}+\varepsilon}$ (robust scale only for standard deviation features)

**Supplementary Note 3: Feature computation**

Optical flow at each time (or frame; 𝑡) and pixel (𝑝) was represented as f_𝑡,𝑝_ = (𝑢_𝑡,𝑝_, 𝑣_𝑡,𝑝_), where 𝑢 and 𝑣 denote the horizontal and vertical flow components, respectively.

**Movement magnitude (mean per-pixel flow displacement):**

$$m_{t,p}=\parallel\mathbf{f}_{t,p}\parallel=\sqrt{u_{t,p}^{2}+v_{t,p}^{2}}$$

At each frame (𝑡), 𝑚_𝑡,𝑝_ was computed for each pixel and summarized using a robust mean. Specifically, values within the 20^th^ to 99.75^th^ percentile range of {𝑚_𝑡,𝑝_}_𝑝_ were retained, and the mean was computed across the resulting pixel magnitudes. The per-frame summaries were then aggregated into non-overlapping 0.5 s bins (5 frames at 10 fps) to smooth the signal.

**Movement velocity (mean change in displacement between consecutive frames):**

Movement velocity was defined as the finite-difference rate of change of the binned motion-magnitude summary. The per-frame magnitude summaries were aggregated into non-overlapping time bins of Δ𝑡 = 0.5 s, and the resulting per-bin mean was denoted as 𝑀_𝑡_. A first-order finite difference was computed using Δ𝑡 = 0.5 s. The first value was set to 𝑉_1_ = 0.

$$V_{t}=\frac{M_{t}-M_{t-1}}{\Delta t}, t=2,\ldots,T$$

**Movement direction (dominant motion direction within a frame):**

The per-pixel motion angle was computed as:

$$\theta_{t,p}=atan2\left( v_{t,p},u_{t,p} \right)\in\left[ -\pi,\pi\right]$$

To account for angular wrap-around at ±𝜋, motion direction was summarized using a circular mean. Unit vectors (cos 𝜃_𝑡,𝑝_, sin 𝜃_𝑡,𝑝_) were summed across pixels and converted back to an angle:

$$\bar{\theta}_{t}=atan2\left( \sum_{p} \sin\left( \theta_{t,p} \right),\sum_{p} \cos\left( \theta_{t,p} \right) \right)$$

$$\bar{\theta}_{t}^{\left( \deg\right)}=\frac{180}{\pi} \bar{\theta}_{t}$$

Bin-level direction was obtained by averaging 𝜃_t_^¯(deg)^ across frames within each Δ𝑡 = 0.5 s bin.

**Movement directional variability (variability of motion angles):**

The dispersion of motion directions within a frame was computed using circular variance, representing movement complexity. The mean resultant length was computed from the summed unit vectors, where 𝑁 was the number of pixels in the frame:

$$R_{t}=\frac{\sqrt{\left( \sum_{p} \sin\left( \theta_{t,p} \right) \right)^{2}+\left( \sum_{p} \cos\left( \theta_{t,p} \right) \right)^{2}}}{N}$$

Directional variability was defined as:

$$\mathrm{Var}_{t}^{\left( \mathrm{ang} \right)}=1-R_{t}$$

Larger values indicate greater dispersion of motion directions within the frame correspond to more dispersed directions within the frame. Bin-level values were obtained by averaging across frames within each Δ𝑡 = 0.5 s bin.

**Movement amplitude (the fraction of pixels exhibiting motion above a threshold):**

For each frame (𝑡), the fraction of pixels whose motion magnitude exceeded a patient-specific threshold 𝜏 was computed as:

$$F_{t}\left( \tau\right)=\frac{1}{HW}\sum_{p=1}^{HW} \mathbb{I}\left[ m_{t,p}>\tau\right]$$

where 𝐻 × 𝑊 denotes frame size and 𝕀[·] represents the indicator function.

**Movement duration (temporal length of a detected movement episode):**

If a clip started at 𝑡_start_ and ended at 𝑡_end_, the duration was calculated as:

$$D=t_{\mathrm{end}}-t_{\mathrm{start}}$$

For each movement feature, additional measures, including percentile values, were computed to characterize within-clip distributions, generating a set of highly correlated ancillary features. However, given the heavily tailed nature of the data, these percentile-based features could be more informative than the mean value itself.

**Supplementary Note 4: Patient-Adaptive Movement Thresholding for Pretraining Clip Extraction**

To account for inter-individual differences in baseline movement activity, patient-specific thresholds were used to segment movement clips for pretraining. For each participant, the mean residual life (MRL) above the 90th percentile of the smoothed optical flow magnitude distribution was computed as a measure of tail heaviness:

$MRL(u) = E[X - u | X > u]$.

A high MRL value indicates a heavy-tailed distribution with frequent large-magnitude movements ("high-MRL" or active sleepers), whereas a low MRL value indicates a more concentrated distribution with fewer large movements ("low-MRL" or calm sleepers).

Based on this measure, two thresholds were applied:

- Low-MRL (calm) sleepers: 97th percentile threshold, to minimize false positives from background noise
- High-MRL (active) sleepers: 93rd percentile threshold, to ensure sufficient capture of true movement events

This patient-adaptive approach allowed the number of retained clips to vary across participants according to their individual activity level, rather than applying a fixed population-level threshold that would systematically over- or under-segment participants at the extremes of the activity distribution. Threshold parameters were tuned through visual inspection in a representative subset of participants spanning the range of activity levels.

Following thresholding, movement clips were extracted using stability rules: a minimum clip duration of 1 second, bridging of short immobility gaps to avoid artificial fragmentation, and hysteresis to prevent repeated triggering at threshold boundaries.

**Supplementary Note 5: Continuous Severity Score Computation**

In the continuous severity setting, clip-level predicted probabilities were converted into point contributions using model-specific scaling functions.

For the SVC model, the predicted probability of the moderate-to-severe class (p) was multiplied by 5, yielding per-clip contributions ranging from 0 to 5: contribution = 5 × p

For the foundation model, continuous point contributions were computed as: contribution = 1 + 4p

This formulation assigned each clip a score between 1 and 5, yielding a lower bound of 1 point consistent with the discrete mild weight and scaling continuously up to the moderate-to-severe weight of 5 when p = 1.

Whole-night severity scores were then calculated as the sum of per-clip contributions across all retained REM movement clips for each patient.

**Supplementary Note 6: Definition of Whole-night Severity Error Metrics**

For each patient 𝑖, 𝑦_𝑖_ and 𝑦̂_𝑖_ denote the true and predicted discrete night scores, respectively, while 𝑦_𝑖_^(𝑐)^ and 𝑦̂_𝑖_^(𝑐)^ denote their continuous counterparts. 𝑛_𝑖_ represents the total number of clips for that night.

Pred (pts) = $\hat{y}_{i}$	Pred (cont) = $\hat{y}_{i}^{\left( c \right)}$

Err (pts) = $\hat{y}_{i}-y_{i}$	|Err| (pts) = $\left| \hat{y}_{i}-y_{i} \right|$	Pct Err = $100\cdot\frac{\left| \hat{y}_{i}-y_{i} \right|}{y_{i}}$

Err (cont) = $\hat{y}_{i}^{\left( c \right)}-y_{i}^{\left( c \right)}$	|Err| (cont) = $\left| \hat{y}_{i}^{\left( c \right)}-y_{i}^{\left( c \right)} \right|$	Pct Err (cont) = $100\cdot\frac{\left| \hat{y}_{i}^{\left( c \right)}-y_{i}^{\left( c \right)} \right|}{y_{i}^{\left( c \right)}}$

|Err| pts/clip = $\frac{\left\vert\hat{y}_{i}-y_{i} \right\vert}{n_{i}}$ Bias pts/clip = $\frac{\hat{y}_{i}-y_{i}}{n_{i}}$

|Err| cont/clip = $\frac{\left\vert\hat{y}_{i}^{\left( c \right)}-y_{i}^{\left( c \right)} \right\vert}{n_{i}}$ Bias cont/clip = $\frac{\hat{y}_{i}^{\left( c \right)}-y_{i}^{\left( c \right)}}{n_{i}}$

Over = $\mathbb{1}\left[ \hat{y}_{i}-y_{i}>0 \right]$	Under = $\mathbb{1}\left[ \hat{y}_{i}-y_{i}<0 \right]$	Exact = $\mathbb{1}\left[ \hat{y}_{i}-y_{i}=0 \right]$

Let 𝑁 denote the number of patients (nights) in the split. For each patient 𝑖, the discrete night-level error was defined as 𝑒_𝑖_ = 𝑦̂_𝑖_ − 𝑦_𝑖_ and the absolute error 𝑎_𝑖_ = |𝑒_𝑖_ |, with per-clip normalization 𝑒_𝑖_^clip^ = 𝑒_𝑖_/𝑛_𝑖_ and 𝑎_𝑖_^clip^ = 𝑎_𝑖_/𝑛_𝑖_. Percent error was calculated as 𝑝_𝑖_ = 100 × 𝑎_𝑖_/𝑦_𝑖_ (%). These formulations were applied identically to continuous scores by substituting discrete values (𝑦_𝑖_, 𝑦̂_𝑖_) with their continuous counterparts (𝑦_𝑖_^(𝑐)^, 𝑦̂_𝑖_^(𝑐)^).

MAE/clip = $\frac{1}{N}\sum_{i=1}^{N} a_{i}^{\mathrm{clip}}$	Median AE/clip = $\mathrm{median}_{i=1..N}\left( a_{i}^{\mathrm{clip}} \right)$	Bias/clip = $\frac{1}{N}\sum_{i=1}^{N} e_{i}^{\mathrm{clip}}$

MAE/night = $\frac{1}{N}\sum_{i=1}^{N} a_{i}$	Median AE/night = $\mathrm{median}_{i=1..N}\left( a_{i} \right)$	Bias/night = $\frac{1}{N}\sum_{i=1}^{N} e_{i}$

Mean % error/night = $\frac{1}{N}\sum_{i=1}^{N} p_{i}$	Median % error/night = $\mathrm{median}_{i=1..N}\left( p_{i} \right)$

|  | iRBD (n = 86) | No RBD (n = 111) | Secondary RBD (n=17) |
| --- | --- | --- | --- |
| *Demographics* | | | |
| Age, mean (SD) | 65.0 (8.6) | 63.8 (9.2) | 71.6 (7.8) |
| Male, n (%) | 68 (79.1) | 79 (71.2) | 12 (70.6%) |
| Healthy sleeper, n (%) | - | 16 (14.4) | - |
| *Sleep disorders* | | | |
| Insomnia, n (%) | 4 (4.7%) | 6 (5.4%) | 5 (29%) |
| RLS, n (%) | 11 (12.8%) | 5 (4.5%) | 4 (23.6%) |
| Sleepwalking, n (%) | 4 (4.7%) | 1 (0.9%) | 2 (11.8%) |
| Sleep talking, n (%) | - | 6 (5.4%) | - |
| Narcolepsy type 1, n (%) | 0 (0) | 2 (1.8%) | 0 (0) |
| AHI3A ≥ 15, n (%) | 21 (24.4%) | 41 (36.9%) | 3 (17.6%) |
| PAP therapy, n (%) | 25 (40.3%) | 43 (48.3%) | 5 (29.4%) |
| *Sleep variables* | | | |
| Total sleep time, min, mean (SD) | 381.4 (78.8) | 361.7 (76.5) | 356.8 (86.5) |
| NREM sleep time, min, mean (SD) | 309.5 (70.0) | 297.6 (68.3) | 288.4 (76.2) |
| REM sleep time, min, mean (SD) | 72.2 (43.7) | 64.8 (44.6) | 68.4 (39.8) |
| AHI3A, mean (SD) | 11.8 (10.9) | 16.7 (14.9) | 16.2 (14.1) |
| REM AHI, mean (SD) | 10.8 (10.8) | 18.5 (14.2) | 14.7 (13.5) |
| PLM index, mean (SD) | 30.7 (36.2) | 14.3 (19.5) | 38.9 (40.7) |
| PLM index ≥ 15, n (%) | 48 (55.8) | 37 (33.3%) | 11 (64.7%) |
| RSWA index, mean (SD) | 0.64 (0.26) | - | 0.70 (0.23) |

**Supplementary Table 1. Demographic, clinical, and polysomographic features of each cohort.** AHI3A, apnea-hypopnea index scored using hypopneas associated with ≥3% oxygen desaturation or arousal; NREM, non-rapid eye movement; OSA, obstructive sleep apnea; PAP, positive airway pressure; PLM, periodic limb movement; REM, rapid eye movement; RLS, restless legs syndrome; RSWA, REM sleep without atonia; SD, standard deviation.

| Feature Type | Split | PCA | Model | Accuracy | Macro F1 | Precision | Recall | AUC |
| --- | --- | --- | --- | --- | --- | --- | --- | --- |
| Raw | **Easy** | **No PCA** | **RF** | 0.74 | 0.56 | 0.57 | 0.68 | 0.74 |
|  |  | **PCA** | **RF** | 0.71 | 0.53 | 0.54 | 0.53 | 0.66 |
|  | **Hard** | **No PCA** | **SVC** | 0.91 | 0.65 | 0.70 | 0.62 | 0.83 |
|  |  | **PCA** | **RF** | 0.84 | 0.60 | 0.59 | 0.63 | 0.73 |
| Normalized | **Easy** | **No PCA** | **XGB** | 0.71 | 0.55 | 0.57 | 0.68 | 0.74 |
|  |  | **PCA** | **RF** | 0.75 | 0.55 | 0.56 | 0.65 | 0.73 |
|  | **Hard** | **No PCA** | **SVC** | 0.90 | 0.55 | 0.61 | 0.54 | 0.79 |
|  |  | **PCA** | **RF** | 0.77 | 0.57 | 0.57 | 0.65 | 0.78 |

**Supplementary Table 2. Heuristic analyses not including movement duration across easy and hard splits.** AUC, area under the curve; PCA, principal component analysis; RF, Random Forest; SVC, Support Vector Classifier; XGB, XGBoost.

| Feature Type | Split | PCA | Model | Accuracy | Macro F1 | Precision | Recall | AUC |
| --- | --- | --- | --- | --- | --- | --- | --- | --- |
| Raw | **Easy** | **No PCA** | **LR** | 0.85 | 0.67 | 0.64 | 0.76 | 0.78 |
|  |  | **PCA** | **SVC** | 0.80 | 0.62 | 0.60 | 0.73 | 0.78 |
|  | **Hard** | **No PCA** | **LR** | 0.85 | 0.65 | 0.63 | 0.71 | 0.84 |
|  |  | **PCA** | **SVC** | 0.83 | 0.66 | 0.63 | 0.75 | 0.82 |
| Normalized | **Easy** | **No PCA** | **LR** | 0.85 | 0.67 | 0.64 | 0.76 | 0.78 |
|  |  | **PCA** | **SVC** | 0.81 | 0.62 | 0.60 | 0.73 | 0.77 |
|  | **Hard** | **No PCA** | **SVC** | 0.81 | 0.62 | 0.61 | 0.71 | 0.81 |
|  |  | **PCA** | **SVC** | 0.82 | 0.65 | 0.62 | 0.75 | 0.81 |

**Supplementary Table 3. Heuristic analyses including movement duration across easy and hard splits.** AUC, area under the curve; LR, Logistic Regression; PCA, principal component analysis; SVC, Support Vector Classifier.

| Strategy | Accuracy | Macro F1 score | Precision | Recall | AUC |
| --- | --- | --- | --- | --- | --- |
| Naïve (Uniform-Frozen-3cls) | 0.88 | 0.40 | 0.38 | 0.42 | 0.65 |
| Uniform Sampling | 0.91 | 0.72 | 0.74 | 0.72 | 0.87 |
| Peak Flow Window (Uniform) | 0.92 | 0.74 | 0.74 | 0.75 | 0.88 |
| Peak Flow Window (Random) | 0.90 | 0.74 | 0.71 | 0.79 | 0.91 |
| Maximum Optical Flow | 0.93 | 0.76 | 0.75 | 0.77 | 0.90 |

**Supplementary Table 4. Performance comparison of frame sampling strategies using V-JEPA2 with checkpoint-based fine-tuning (Easy split).** The naïve baseline used uniform frame sampling with a frozen encoder and three-class output (mild, moderate, and severe). Uniform sampling selected frames at equal intervals across the clip. Peak-flow window strategies selected frames uniformly or randomly within the 3-second temporal window of highest optical flow activity. Maximum optical flow selected the 16 frames with the highest optical flow magnitudes across the clip. Positive class is moderate-to-severe movements. AUC, area under the curve; 3cls, three-class classification. AUC, area under the curve.

| Strategy | Accuracy | Macro F1 | Precision | Recall | AUC |
| --- | --- | --- | --- | --- | --- |
| Frozen Encoder | 0.69 | 0.49 | 0.52 | 0.57 | 0.68 |
| Unfrozen Encoder | 0.88 | 0.69 | 0.67 | 0.78 | 0.82 |
| Two-Stage Fine-Tuning  (Peak Flow uniform) | 0.89 | 0.71 | 0.67 | 0.78 | 0.83 |
| Two-Stage Fine-Tuning  (Maximum Optical Flow) | 0.90 | 0.73 | 0.69 | 0.80 | 0.86 |

**Supplementary Table 5. Comparison of fine-tuning strategies using custom domain-specific pretraining (Easy split).** AUC, area under the curve.

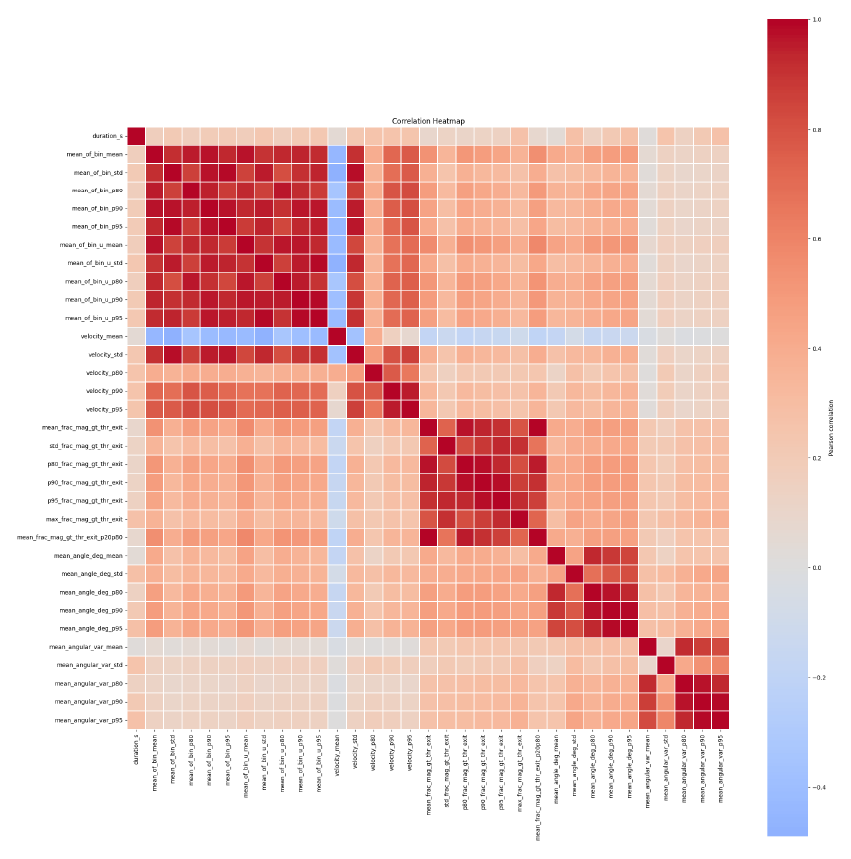

**Supplementary Figure 1. Correlation heatmap of extracted movement features before correlation-threshold filtering.** Pairwise Pearson correlation coefficients are shown for all derived features. Warmer colors indicate positive correlations, cooler colors indicate negative correlations, and white indicates near-zero correlations.

duration_s, mean_of_bin_mean, mean_of_bin_std, mean_of_bin_p80, mean_of_bin_p90, mean_of_bin_p95, mean_of_bin_u_mean, mean_of_bin_u_std, mean_of_bin_u_p80, mean_of_bin_u_p90, mean_of_bin_u_p95, velocity_mean, velocity_std, velocity_p80, velocity_p90, velocity_p95, mean_frac_mag_gt_thr_exit, std_frac_mag_gt_thr_exit, p80_frac_mag_gt_thr_exit, p90_frac_mag_gt_thr_exit, p95_frac_mag_gt_thr_exit, max_frac_mag_gt_thr_exit, mean_frac_mag_gt_thr_exit_p20p80, mean_angle_deg_mean, mean_angle_deg_std, mean_angle_deg_p80, mean_angle_deg_p90, mean_angle_deg_p95, mean_angular_var_mean, mean_angular_var_std, mean_angular_var_p80, mean_angular_var_p90, mean_angular_var_p95

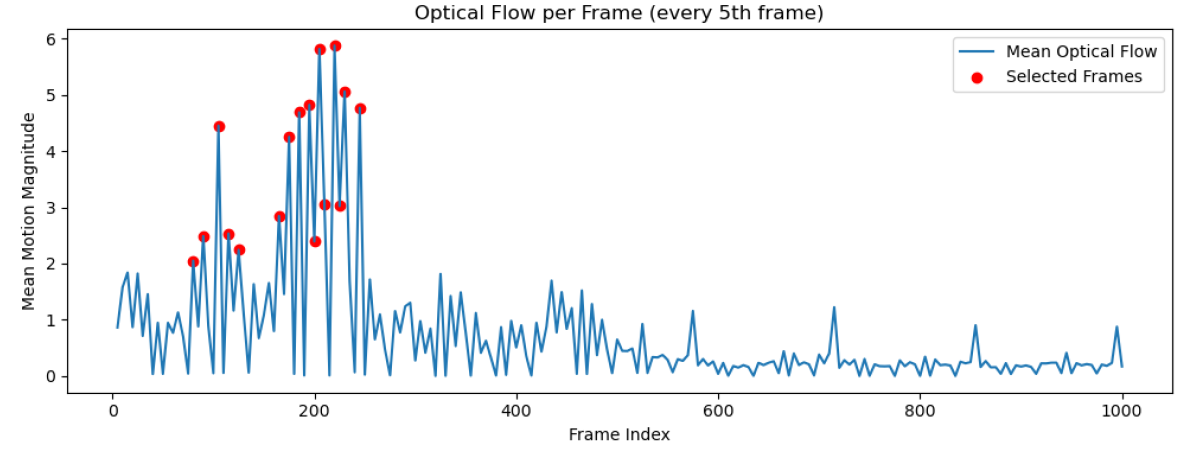

**Supplementary Figure 2. Illustration of maximum optical flow-based frame sampling for a representative movement clip.**

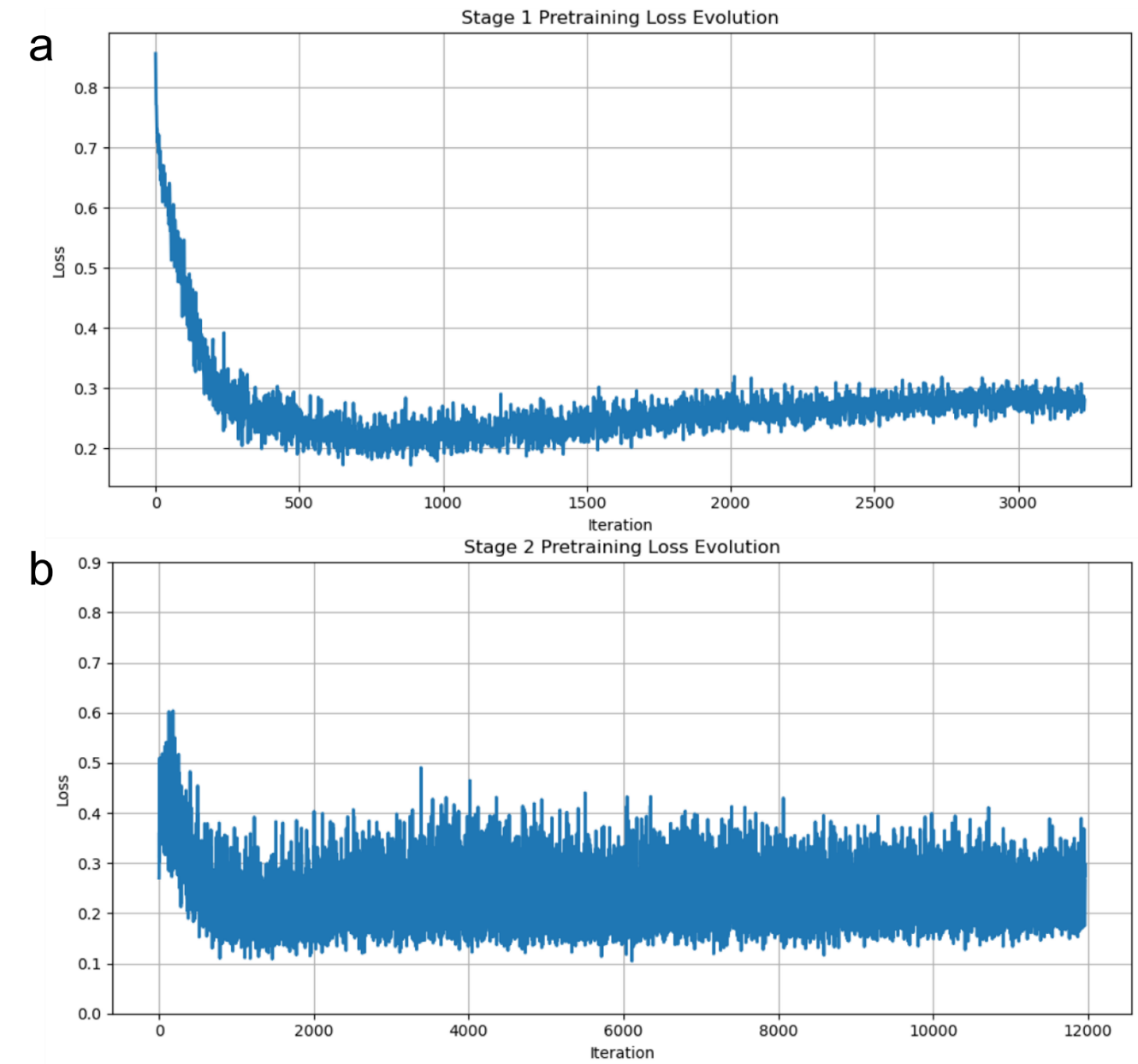

**Supplementary Figure 3. Original V-JEPA2 pretraining losses per iteration.**

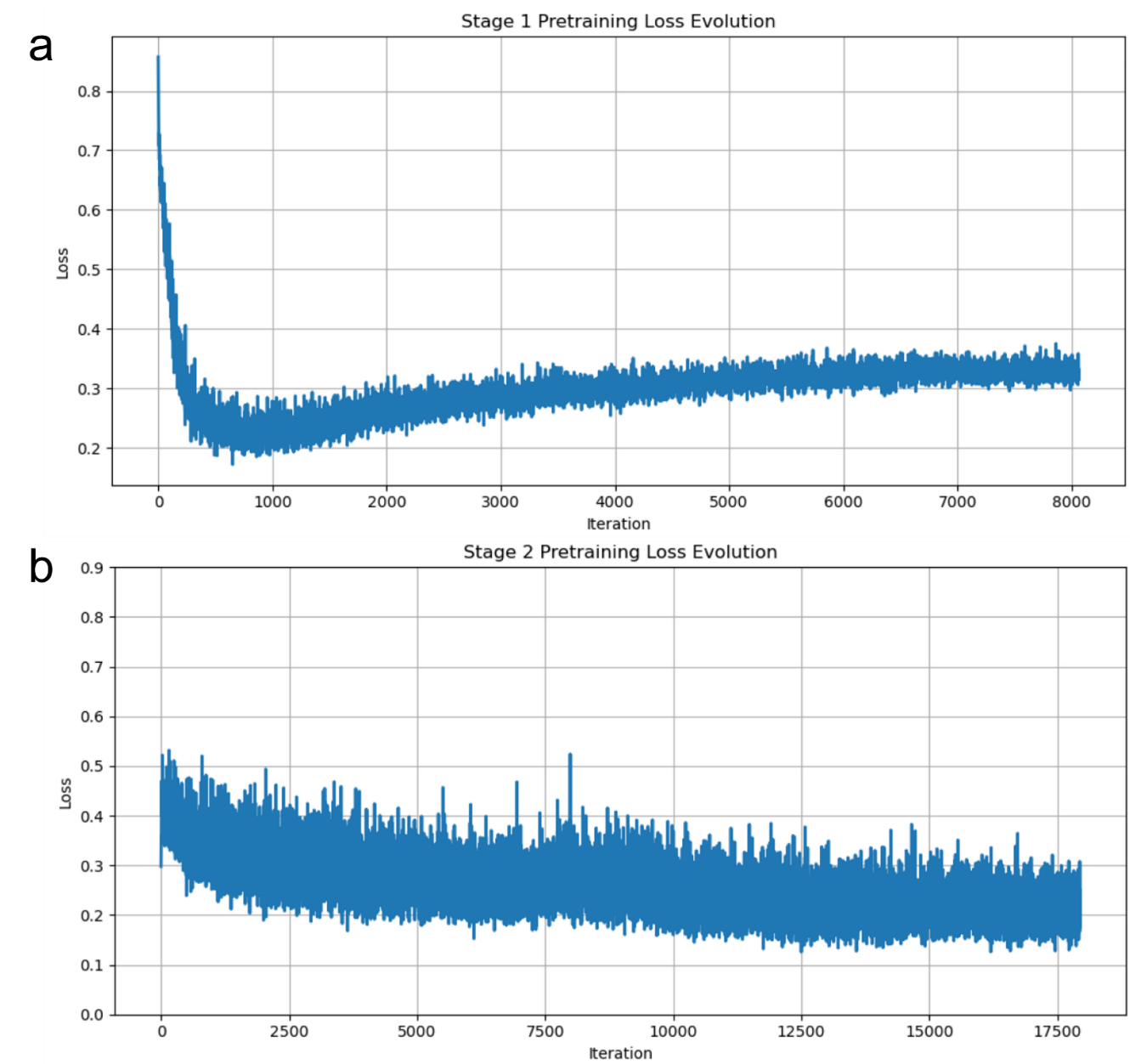

**Supplementary Figure 4. Scaled two-stage V-JEPA2 pretraining losses per iteration.**

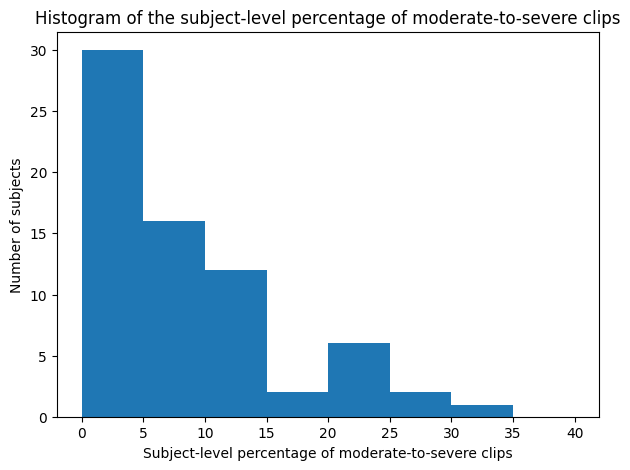

**Supplementary Figure 5. Histogram showing the distribution of the subject-level percentages of moderate-to-severe movement clips.**

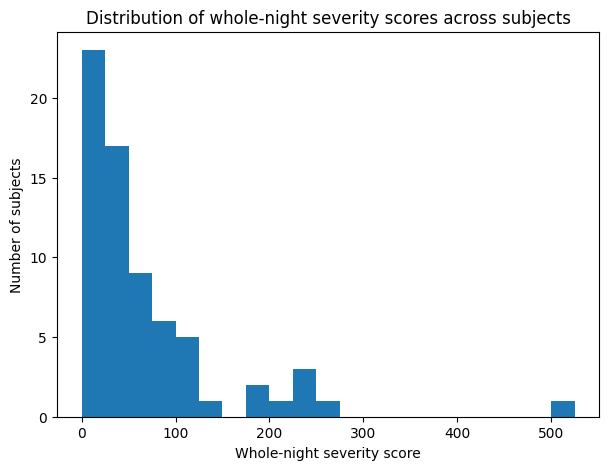

**Supplementary Figure 6. Distribution of whole-night severity scores across subjects, with clip-level severity labels weighted as 1 for mild and 5 for moderate-to-severe.**
